# Seroprevalence and awareness of hepatitis C infection among informal workers at Kejetia Market, Ghana: A cross-sectional study

**DOI:** 10.64898/2026.10.04.26364720

**Authors:** Daniel Kobina Okwan, De-Graft Kwaku Ofosu Boateng, Pius Takyi, Michael Agyemang Obeng, Joshua Kofi Attah, Patrick Oduro, Clinton Owusu Boateng, Senam Yawa Nunamey Ahadzie, Godfred Yawson Scott, Augustine Yeboah, Akwasi Amponsah Abrampah, Kwaku Duah Ibrahim, Felix Kusi Boakye, Daniel Dzemedo Nouwati, Kofi Bonsu Amankwah, Abdul Latif Koney Shardow, Abu Abudu Rahamani

## Abstract

Globally, Hepatitis C Virus (HCV) infection is a leading cause of chronic liver disease and remains a significant public health threat in Ghana. However, there is a paucity of data on the prevalence and awareness of HCV among market sellers and informal workers in Ghana. This study sought to determine the prevalence and awareness of HCV among this population.

This was a cross-sectional study conducted among 397 sellers and workers at Kejetia market from December 2024 to January 2025. HCV was diagnosed using the One Test Hepatitis C Virus Test Strip (Qingdao Hightop) for the qualitative detection of HCV antibodies. Data were analysed using SPSS version 26 and GraphPad Prism version 8.0. A *p < 0.05* was considered statistically significant for awareness of HCV.

The prevalence of HCV in this study was 4 (1.0%). Awareness of HCV was low at 112 (28.2%). Residence (*p*=0.021) and age group (*p*=0.040) were significantly associated with HCV awareness. Age 41-50 years was almost three times more likely to be aware of Hepatitis C infection compared to ≤ 30 years.

Although the prevalence of HCV was low, awareness of the infection was also limited; this highlights the need for public health authorities to implement targeted HCV education programs in major markets to effectively reach informal sector workers who are often overlooked by conventional campaigns.

## Introduction

Hepatitis C virus (HCV) infection is a blood-borne disease transmitted through direct exposure to infected blood, such as via blood transfusions, or through exposure to contaminated instruments during unsafe injections, medical procedures, or other percutaneous practices (1). Globally, HCV is a leading cause of chronic liver disease, with persistent HCV infection strongly linked to cirrhosis, hepatocellular carcinoma, and liver cancer (2). HCV is a small, single-stranded, enveloped RNA virus belonging to the family *Flaviviridae* and exhibits a high degree of genetic heterogeneity. Seven distinct HCV genotypes have been identified, of which genotype 1 is the most prevalent worldwide, accounting for approximately 46% of all infections(3).

The World Health Organization (WHO) estimated that 57.8 million people were living with chronic HCV infection worldwide in 2019. During the same year, more than 3 million people acquired chronic hepatitis B virus (HBV) or HCV infection, including approximately 1.5 million new chronic HCV infections, with the WHO African Region accounting for 39.6% of all new chronic HBV and HCV infections (4). Furthermore, an estimated 300,000 HCV-related deaths occur annually. Despite these figures, the true burden of HCV is likely underestimated because the infection is frequently asymptomatic for many years and may present with extrahepatic manifestations that delay diagnosis (5).

HCV infection is the second leading cause of end-stage liver disease and hepatocellular carcinoma-related mortality in sub-Saharan Africa. The overall HCV seroprevalence in the region is estimated at 2.98% (2). In Ghana, the national prevalence of HCV is estimated to be 3.0%, placing the country among those with a relatively high burden of infection in the region (6). Similarly, a 2016 systematic review comprising hospital-based and limited community-based studies reported an HCV antibody prevalence of 3.0% in Ghana. Within the Ashanti Region, however, the pooled prevalence is comparatively lower (1.5%), based on 13 studies involving 87,805 participants (7).

Elle Saine *et al*. (2021) reported widespread misconceptions and substantial knowledge gaps regarding HCV among infected individuals. Limited awareness and knowledge of HCV have been identified as major barriers to the uptake of HCV testing by patients and the provision of testing by healthcare providers. Consequently, poor awareness contributes to ongoing transmission and missed opportunities for prevention, early diagnosis, and treatment (9). Because there is currently no effective vaccine against HCV, the WHO has prioritized increasing public awareness, expanding screening services, and improving access to care and treatment as key strategies toward HCV elimination. Adequate knowledge of HCV is therefore essential for preventing transmission and encouraging individuals to seek timely testing, care, and treatment (10).

Most previous studies on HCV in Ghana have been hospital-based, limiting their generalizability to the wider community and understudied occupational groups. Consequently, there is limited evidence on the burden of HCV among informal workers and market sellers, despite their potential vulnerability to infection. In addition to the lack of prevalence data, information on HCV awareness within this population remains scarce. As reported by Obeng *et al.* (2026), informal sector workers may be particularly vulnerable to HCV because of limited access to occupational health services, irregular income, and reduced exposure to health education and screening programmes.

In line with global efforts, the WHO has set a target to eliminate viral hepatitis as a public health threat by 2030, emphasizing the need to identify high-risk populations and improve awareness through targeted interventions (12, 13). Achieving this target requires locally generated evidence to inform context-specific public health policies and interventions, particularly among underserved populations.

Kejetia Market, one of the largest commercial centres in Ghana, accommodates thousands of traders and informal workers who may unknowingly be exposed to HCV risk factors. However, data on HCV prevalence and awareness in this population are lacking. Therefore, this study aimed to determine the seroprevalence of HCV infection and assess awareness of HCV among informal workers at Kejetia Market, Kumasi, Ghana. The findings will provide evidence to guide targeted health education, screening programmes, and other public health interventions for informal sector workers and contribute to Ghana’s efforts toward achieving the WHO target of eliminating viral hepatitis as a public health threat by 2030 (14).

## Materials and methods

### Study design and setting

A cross-sectional study was conducted to assess the prevalence of HCV infection among the sellers and workers at Kejetia Market, Ghana. The cross-sectional design was selected to provide a representation of the HCV prevalence and associated risk factors within this occupational group in Ghana. The study was conducted from 8^th^ June 2024 to 5^th^ January 2025 at the Kejetia Market in Kumasi, one of the largest commercial centres in Ghana, with thousands of traders and workers. Kumasi is the second largest city in Ghana, situated between latitudes 5.35°N and 6.40°N and longitudes 1.3°W and 1.35°W. It covers an approximate area of 150km^2^, residing within the rainforest region of West Africa. The city has a population of about two million inhabitants and is suitable for carrying out this study due to its cosmopolitan status(13). The Kejetia Market, also known as the Kumasi Central Market, has three floors, with most of the facilities - banks, clinics, transport systems, butcher’s yards, police stations, and the administration block - on the ground floor. The first and second floors are similar -- in terms of features, with restaurants, stores, and tailor’s yards. The market covers 172,197m^2^ with over 10,000 stores and stalls, with an average temperature of 24°C and humidity of 85%(15).

### Sample size and sampling method

The sample size was determined using Cochran’s formula for prevalence studies using an estimated HCV prevalence rate of 14.4%(16) at a 95% confidence level and a 5% margin of error.

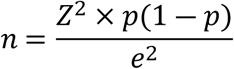

Where: n = required sample size, Z = Z-score at 95% confidence level (1.96), p = estimated prevalence (0.144), 1 − p = 0.856, e = margin of error (0.05).

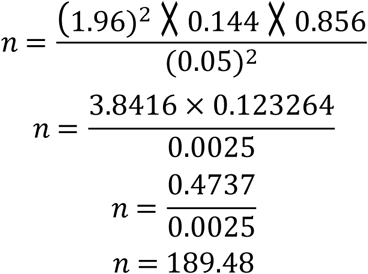

The minimum sample size required was 190. However, to improve statistical power, 397 participants were recruited for this study. A stratefied random sampling technique was employed to recruit participants from different sections of the Kejetia Market. To guarantee effective representation across various market zones, the market was divided into sections, and participants were randomly chosen from each zone. In collaboration with the leadership of the market and the registered trader associations, which keep records of traders and workers across the different market sections, the sampling frame was developed. An estimated population size of each association was determined from these records and validated by leaders of the associations. Across all associations, every 5^th^ individual on the list was selected to be included in the study. This structured approach effectively reduced selection bias and ensured that the participants of the study reflected the occupational composition of informal sector workers at the Kejetia Market.

### Study population, inclusion, and exclusion criteria

The target population for this study comprised sellers and workers employed by the market management, aged 18 years and above, who were actively engaged in commercial activities at Kejetia. The sellers recruited for this study included butchers and meat sellers, fishmongers, food vendors, general goods store keepers, mobile workers (hawkers), cloth and textile workers. The workers recruited for this study included barbers and hairdressers, tailors and seamstresses, waste collectors and cleaners, market porters, as well as drivers and mates. However, for both sellers and workers, only individuals aged 18 years or older and were willing to provide informed consent to the study were included in this study. Individuals who were unwilling to participate in the study were excluded from the study.

### Data collection

A well-structured questionnaire was validated and administered to the participants within the framework of the study variables, consisting of participant demographics and other information related to the risk factors and awareness of HCV. The data were collected using an investigator-administered questionnaire in a language that they could easily comprehend (English and Twi). All researchers involved in the study underwent training before the start of the study.

After administering the questionnaire and obtaining all necessary information, the participant’s ring finger was gently massaged to increase blood flow to the tip of the finger and then disinfected with an alcohol swab. Afterwards, a sterile lancet was used to prick the finger, and about 10µl of blood was collected onto the One Test Hepatitis C Virus Test Strip (Qingdao Hightop Biotech Co. Ltd., China) for the qualitative detection of HCV antigens. The test strips were equilibrated and tested at room temperature before the commencement of the study. After the blood sample was added to the test strip, a drop of HCV buffer was added, and the results were recorded after 10 minutes. The appearance of two separate lines in the control and test regions, respectively, indicated a positive result, whereas a single red line in the control region, without any visible line in the test region, indicated a negative result. If no red line showed in the control region, the test was considered invalid and repeated.

### Data analysis

The primary outcome variable was HCV seropositivity, which was determined through serological testing. Independent variables included age, sex, occupation, history of blood transfusions, sharing of sharp instruments, and awareness of HCV. Awareness of HCV was binary and was coded as (Yes: 1, No:0). HCV infection was also binary and was coded as (Positive: 1, Negative:0). Data were analysed using SPSS version 26 and GraphPad Prism version 8.0. Descriptive statistics were used to summarize participant characteristics. Chi-square tests and logistic regression were employed to determine the associations between risk factors and HCV prevalence. A p-value <0.05 was considered statistically significant.

### Ethical considerations

This study was conducted in accordance with the principles of the Declaration of Helsinki. Ethical approval for this study was obtained from the Committee on Human Research, Publication, and Ethics (CHRPE), School of Medical Sciences, Kwame Nkrumah University of Science and Technology (KNUST), with approval number (CHRPE/AP/180/24).

An approval letter was also obtained from the management of the Kejetia Market before the commencement of the study. All participants and leaders of the workers’ association at the market gave their written informed consent after the aim and procedure of the study were explained to them. All data collected were anonymized to ensure confidentiality, and participation was voluntary with the right to withdraw from the study at any stage without penalty. Written informed consent was obtained from all participants prior to recruitment. Participants with positive results were informed confidentially and referred to the Komfo Anokye Teaching Hospital (KATH) for a confirmatory HCV infection (PCR) testing.

## Results

### Background characteristics of study participants

Most (40.3%) of the participants in this study were ≤ 30 years, were female 252 (63.5%), were married 209 (52.6%), and were Christians 265 (66.8%). The majority of them had basic education 183 (46.1%), lived in urban areas 286 (72.0%), and were market porters 68 (17.1%). Most 271 (68.3%) of the study participants have been selling at Kejetia for ≤ 10 years (**Table 1**)

**Table 1.** Background characteristics of the study participants.

| Variables | Frequency (n=397) | Percentage (%) |
| --- | --- | --- |
| **Age Group** |  |  |
| $\leq 30$ years | 160 | 40.3 |
| 31-40 years | 74 | 18.6 |
| 41-50 years | 85 | 21.4 |
| $\geq 51$ years | 78 | 19.6 |
| **Gender** |  |  |
| Female | 252 | 63.5 |
| Male | 145 | 36.5 |
| **Marital Status** |  |  |
| Co-Habiting | 40 | 10.1 |
| Divorced | 13 | 3.3 |
| Married | 209 | 52.6 |
| Single | 115 | 29.0 |
| Widowed | 20 | 5.0 |
| **Religion** |  |  |
| Christian | 265 | 66.8 |
| Islamic | 125 | 31.5 |
| Non-Religious | 7 | 1.8 |
| **Educational Level** |  |  |
| No Education | 47 | 11.8 |
| Basic | 183 | 46.1 |
| Secondary | 133 | 33.5 |
| Tertiary | 34 | 8.6 |
| **Residence** |  |  |
| Rural | 111 | 28.0 |
| Urban | 286 | 72.0 |
| **Work Type** |  |  |
| Butchers and meat sellers | 46 | 11.6 |
| Drivers and mates | 29 | 7.3 |
| Fish mongers | 44 | 11.1 |
| Food vendors | 43 | 10.8 |
| General goods store keeper | 55 | 13.9 |
| Market porters (Kayaye) | 68 | 17.1 |
| Mobile workers (Hawkers) | 47 | 11.8 |
| Tailors and seamstress | 53 | 13.4 |
| Waste collectors and cleaners | 12 | 3.0 |
| **Duration of stay at Kejetia** |  |  |
| ≤ 10 years | 271 | 68.3 |
| > 10 years | 126 | 31.7 |

Data is presented as frequency (%)

### Prevalence of Hepatitis C infection among study participants

Fig 1 shows the overall prevalence of Hepatitis C among the study participants. Out of the 397 participants recruited for this study, 4 (1.0%) of them tested positive for Hepatitis C infection (Fig 1).

**Figure 1.**
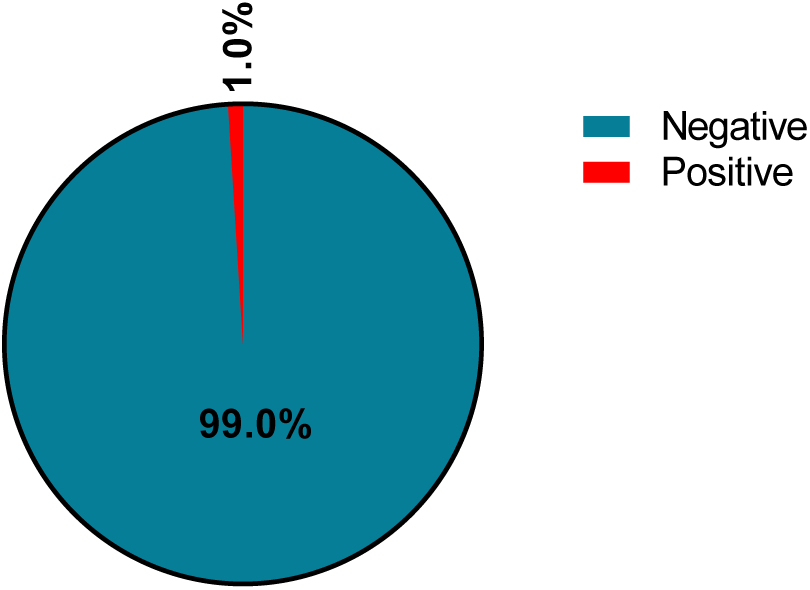
Prevalence of Hepatitis C infection among the study participants.

### Awareness and behavioural characteristics among study participants

Awareness of Hepatitis C infection was measured through a mixed binary and self-rated ordinal scale, with participants responding either yes or no to an awareness of the infection and then rating their knowledge of the infection if aware. The majority 285 (71.8%) of the study participants have not heard about Hepatitis C infection before (Fig 2). The majority of the study participants have multiple sexual partners 262 (66.0%) and have not had STDs before 305 (76.8%). Most 263 (66.2%) of the participants had their first sexual intercourse at ≥ 18 years. Majority of the participants have not been transfused with blood before 344 (86.6%), do not have tattoo 365 (91.9%), and have not injected intravenous drug before 386 (97.2%). 278 (70.0%) of the participants have had their finger/toe nails clipped by nail clippers before (**Table 2**).

**Figure 2.**
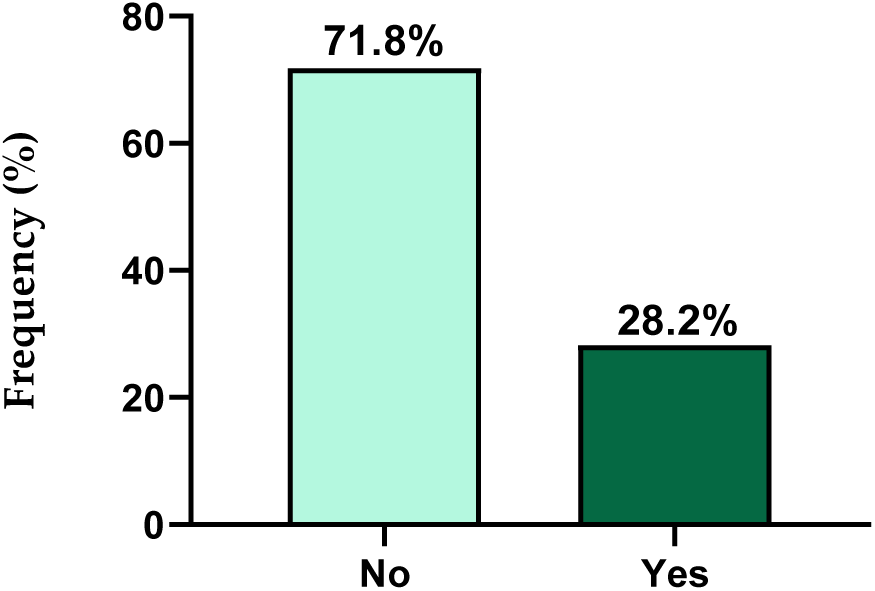
Awareness of Hepatitis C Infection Among Study Participants.

**Table 2.** Behavioural Characteristics of the study participants.

| Variables | Frequency (n=397) | Percentage (%) |
| --- | --- | --- |
| **Number of sexual partners** |  |  |
| Multiple | 262 | 66.0 |
| Single/None | 135 | 34.0 |
| **Have had STDs before** |  |  |
| No | 305 | 76.8 |
| Yes | 92 | 23.2 |
| **Age of first sexual intercourse** |  |  |
| < 18 years | 92 | 23.2 |
| ≥ 18 years | 263 | 66.2 |
| Unwilling to disclose | 42 | 10.6 |
| **Have had blood transfusion before** |  |  |
| No | 344 | 86.6 |
| Yes | 53 | 13.4 |
| **Have tattoo** |  |  |
| No | 365 | 91.9 |
| Yes | 32 | 8.1 |
| **Have had finger/toe nails clipped by nail clippers before** |  |  |
| No | 119 | 30.0 |
| Yes | 278 | 70.0 |
| **Have injected intravenous drug before** |  |  |
| No | 386 | 97.2 |
| Yes | 11 | 2.8 |

Data is presented as frequency (%)

### Association between sociodemographic characteristics and awareness of Hepatitis C infection

In this study, gender (*p*=0.238), marital status (*p*=0.273), religion (*p*=0.520), and educational level (*p*=0.340) were not significantly associated with Hepatitis C infection awareness. However, age group (*p*=0.040) and residence (*p*=0.021) were significantly associated with Hepatitis C awareness (**Table 3**).

**Table 3.** Association between sociodemographic characteristics and Awareness of Hepatitis C infection.

| Variables | Hepatitis C infection awareness |  | *p*-value |
| --- | --- | --- | --- |
|  | No (n=285) | Yes (n=112) |  |
| **Age Group** |  |  | **0.040** |
| ≤ 30 years | 127(79.4) | 33(20.6) |  |
| 31-40 years | 50(67.6) | 24(32.4) |  |
| 41-50 years | 54(63.5) | 31(36.5) |  |
| ≥ 51 years | 54(69.2) | 24(30.8) |  |
| **Gender** |  |  | 0.238 |
| Female | 186(73.8) | 66(26.2) |  |
| Male | 99(68.3) | 46(31.7) |  |
| **Marital Status** |  |  | 0.273 |
| Co-Habiting | 28(70.0) | 12(30.0) |  |
| Divorced | 9(69.2) | 4(30.8) |  |
| Married | 142(67.9) | 67(32.1) |  |
| Single | 89(77.4) | 26(22.6) |  |
| Widowed | 17(85.0) | 3(15.0) |  |
| **Religion** |  |  | 0.520 |
| Christian | 195(73.6) | 70(26.4) |  |
| Islamic | 85(68.0) | 40(32.0) |  |
| Non-Religious | 5(71.4) | 2(28.6) |  |
| **Educational Level** |  |  | 0.340 |
| No Education | 35(74.5) | 12(25.5) |  |
| Basic | 135(73.8) | 47(26.8) |  |
| Secondary | 95(71.4) | 38(28.6) |  |
| Tertiary | 20(58.8) | 14(41.2) |  |
| **Residence** |  |  | 0.021 |
| Rural | 89(80.2) | 22(19.8) |  |
| Urban | 196(68.5) | 90(31.5) |  |

Data is presented as frequency (%). $p < 0.05$ was considered statistically significant for hepatitis C infection awareness.

### Univariate and multivariate logistic regression model of sociodemographic characteristics as predictors of awareness of Hepatitis C infection

In a univariate logistic regression model, age group and residence were the predictors of Hepatitis C infection awareness. Specifically, 41-50 years [cOR= 2.209, 95% CI (1.231-3.964), *p*=0.008] were 2.2 times more likely to be aware of Hepatitis C infection compared to ≤ 30 years. Urban residents [cOR= 1.858, 95% CI (1.094-3.153), *p*=0.022] were almost 2 times more likely to be aware of Hepatitis C infection compared with rural residents. In the multivariate logistic regression model, individuals aged 41-50 years [aOR= 2.427, 95% CI (1.278-4.610), *p*=0.007] were about 2.4 times more likely to be aware of Hepatitis C infection compared to ≤ 30 years (**Table 4**). Due to the low prevalence of HCV infection among the participants, risk factors such as sexual behaviour, transfusion history, and intravenous (IV) drug use were not analyzed to determine their significant correlation with HCV infection.

**Table 4.** Univariate and multivariate logistic regression model of sociodemographic characteristics as predictors of awareness of Hepatitis C infection.

| Variable | cOR | *p*-value | aOR | *p*-value |
| --- | --- | --- | --- | --- |
| **Age Group** |  |  |  |  |
| $\leq 30$ years | 1.00 | | | |
| 31-40 years | 1.847(0.994-3.432) | 0.052 | 1.813(0.955-3.443) | 0.069 |
| 41-50 years | 2.209(1.231-3.964) | **0.008** | 2.427(1.278-4.610) | **0.007** |
| $\geq 51$ years | 1.710(0.925-3.163) | 0.087 | 1.700(0.885-3.264) | 0.111 |
| **Residence** |  |  |  |  |
| Rural | 1.00 |  |  |  |
| Urban | 1.858(1.094-3.153) | **0.022** | 0.662(0.379-1.154) | 0.146 |

cOR, Crude Odds ratio; aOR, Adjusted Odds ratio; CI, Confidence Interval; 1.00, reference.

Binary logistic regression analysis was performed to obtain odds ratios. A p-value of < 0.05 was considered statistically significant. The boldened values indicate p-values that are statistically significant.

## Discussion

This study examined the prevalence of HCV infection among sellers and workers at Kejetia Market, Ghana’s largest commercial centre. The overall prevalence of HCV infection in the study population was 1.0% (95% CI: 0.3%–2.6%) (Figure 1), which falls within the 1-3% prevalence rate in Ghana (17, 18). The prevalence in this study is comparable to the global prevalence of viraemic HCV, estimated at 1·0% in a modelling study (14). However, it is lower than the pooled prevalence of 1.7% reported in the United States (19) and the 2.98% pooled prevalence reported in Sub-Saharan Africa (20). Okonkwo, U. C., *et al.* (2017) (21) has also reported a prevalence of 10.0% in Nigeria.

In Ghana, higher prevalence rates have been reported in previous studies. A systematic review and meta-analysis from 2016-2025 (22) estimated the national pooled prevalence of HCV infection at 6.04%, while another retrospective cross-sectional study reported a prevalence of 1.9% (23). The variations in the prevalences could be due to differences in study populations, geographical settings, and levels of exposure to HCV risk factors, as well as variations in screening practices, preventive interventions over time, and differences in the diagnostic methods used across studies. For instance, Okonkwo, U. C., *et al.* (21) used an enzyme-linked immunosorbent assay (ELISA) for the diagnosis of HCV, which is more accurate and sensitive (24) compared to the point-of-care testing (POCT) used in this study.

Awareness of HCV in this study was low, at 28.2%. This rate is low compared to the 53.74% reported in China (25), 58% in Nigeria (26), and 38.7% in another study in Ghana (27). The relatively low awareness observed in this study may be attributed to the fact that the participants work in the informal sector, a population group that is often overlooked in public health advocacy and awareness campaigns (28, 29). Additionally, many of the participants have had only basic or secondary levels of education, which may limit exposure to targeted health education initiatives (29, 30) on HCV. This emphasizes the need to strengthen targeted health education and awareness campaigns on HCV, particularly among informal sector workers and populations with lower levels of formal education.

In Ghana, HCV transmission is mainly linked to healthcare-related exposures and illicit drug use (31, 32). Interestingly, the majority of participants in this study reported no history of intravenous drug use (97.2%) and no tattoos (91.9%), suggesting that these common risk factors were relatively uncommon within the study population, even though awareness of HCV was low in this study. This is encouraging, as the low prevalence of these risk behaviours could contribute to efforts aimed at achieving the WHO goal of eliminating viral hepatitis, particularly chronic hepatitis B and C, by 2030 (18, 33).

Age group was significantly associated with awareness of HCV in this study, as those aged 41-50 years were almost three times more likely to be aware of Hepatitis C infection compared to ≤ 30 years. This association is true because individuals in older age groups are more likely to have greater life experience, increased exposure to health information, and more frequent interactions with healthcare services through routine medical visits or screening programs, which could increase their awareness and understanding of hepatitis infections. These findings are consistent with the findings in China (25).

In this study, residence was significantly associated with HCV awareness (Table 3.0) but could not independently predict awareness in the multivariate logistic regression analysis (Table 4.0). This is consistent with the findings in the US (34) and China (25). Evidence from literature indicates that residence is a significant social determinant of health, largely due to differences in access to healthcare services, health information, and public health campaigns across residential settings (35–37). Individuals living in urban areas are more likely to be exposed to health education programs, media campaigns, healthcare facilities, and screening services, which can improve their awareness of HCV. In contrast, those residing in rural or less developed areas often have limited access to health information and healthcare services, which may contribute to lower levels of awareness (35–37).

## Conclusion

The prevalence of HCV in this study was 1.0%. Awareness of HCV was low at 28.2%. Residence (*p*=0.021) and age group (*p*=0.040) were significantly associated with HCV awareness. Individuals aged 41-50 years were almost three times as likely to be aware of Hepatitis C infection compared to those aged ≤ 30 years. There was a low occurrence of high-risk behaviors associated with HCV transmission, such as intravenous drug use (2.8%) and tattooing (8.1%), among the study participants. Despite the relatively low prevalence and risk behaviours observed, the limited level of awareness highlights an important gap in public health education. Therefore, public health authorities should design and implement targeted education programs in major markets such as Kejetia to improve knowledge of Hepatitis C, particularly among informal sector workers who are often excluded from conventional health promotion campaigns.

## Strengths and limitations

While the use of rapid diagnostic tests (RDTs) may be practical in field settings, they may have lower sensitivity and specificity compared with ELISA or other molecular assays. Nonetheless, the findings provide valuable insight into HCV epidemiology in a relevant urban working population and contribute to the evidence base guiding HCV prevention and control strategies in Ghana. These findings provide evidence to guide targeted interventions in informal sector populations, which are often underrepresented in surveillance data. Also, systematic sampling improves representation. Furthermore, due to the low HCV seropositivity observed in this study, we were unable to assess the association between independent variables and HCV status or identify its predictors.

## Data Availability

Data is available from the corresponding author upon reasonable request.

## Acknowledgments

We express our sincere appreciation to the management of Kejetia Market and to all the study participants who willingly took part in this study.

## References

1. Roudot-Thoraval F. Epidemiology of hepatitis C virus infection. Clinics and research in hepatology and gastroenterology. 2021;45(3):101596.

2. Sonderup MW, Afihene M, Ally R, Apica B, Awuku Y, Cunha L, et al. Hepatitis C in sub-Saharan Africa: the current status and recommendations for achieving elimination by 2030. The Lancet Gastroenterology & Hepatology. 2017;2(12):910–9.

3. Schillie S. CDC recommendations for hepatitis C screening among adults—United States, 2020. MMWR Recommendations and Reports. 2020;69.

4. Cui F, Blach S, Mingiedi CM, Gonzalez MA, Alaama AS, Mozalevskis A, et al. Global reporting of progress towards elimination of hepatitis B and hepatitis C. The lancet Gastroenterology & hepatology. 2023;8(4):332–42.

5. Lo Moro G, Scaioli G, Vola L, Guastavigna L, Frattin R, De Vito E, et al. Exploring knowledge and awareness of HCV infection and screening test: a cross-sectional survey among an Italian sample. Journal of Community Health. 2023;48(5):769–83.

6. Cofie YND, Senoo-Dogbey VE, Kennedy BTC, Tetteh MD, Mensah MM. Intentions for the prevention of hepatitis B and C among healthcare workers in a tertiary facility in Ghana. IJID regions. 2025;14:100590.

7. Agyeman AA, Ofori-Asenso R, Mprah A, Ashiagbor G. Epidemiology of hepatitis C virus in Ghana: a systematic review and meta-analysis. BMC infectious diseases. 2016;16:1–14.

8. Saine ME, Szymczak JE, Moore TM, Bamford LP, Barg FK, Forde KA, et al. The impact of disease-related knowledge on perceptions of stigma among patients with Hepatitis C Virus (HCV) infection. PLoS One. 2021;16(10):e0258143.

9. Ha S, Timmerman K. Awareness and knowledge of hepatitis C among health care providers and the public: A scoping review. Canada Communicable Disease Report. 2018;44(7-8):157.

10. Liu Y, Su J, Wang X, Xu H, Wang H, Kang R, et al. Hepatitis C Knowledge and Self-Reported Testing Behavior in the General Population in China: Online Cross-Sectional Survey. JMIR Public Health and Surveillance. 2023;9:e39472.

11. Obeng MA, Okwan DK, Boateng CO, Scott GY, Attah JK, Ahadzie SYN, et al. Towards hepatitis B elimination in Ghana: vaccination coverage and its predictors among informal sector workers in Kejetia, Kumasi, Ghana. PloS one. 2026;21(4):e0334205.

12. Obeng MA, Okwan DK, Adankwah E, Owusu PK, Gyamerah SA, Duah KB, et al. Seroconversion and prevalence of Hepatitis B surface antigen among vaccinated health care workers in Ashanti Region, Ghana. Advances in Medicine. 2023;2023(1):2487837.

13. Okwan DK, Scott GY, Takyi P, Boateng CO, Antwi PB, Abrampah AA, et al. A Multicentre Cross-Sectional Study on Hepatitis B Vaccination Coverage and Associated Factors Among Personnel Working in Health Facilities in Kumasi, Ghana. BioMed Research International. 2024;2024(1):8899638.

14. Blach S, Terrault NA, Tacke F, Gamkrelidze I, Craxi A, Tanaka J, et al. Global change in hepatitis C virus prevalence and cascade of care between 2015 and 2020: a modelling study. The Lancet Gastroenterology & Hepatology. 2022;7(5):396–415.

15. Sackey LN, Agyemang LS, Acheampong PE, Owusu MA, Amoah J. Assessment of Noise Levels and Perceptions of Its Health Impact at Kejetia Market in Ghana. The Scientific World Journal. 2024;2024(1):7658837.

16. Nartey YA, Okine R, Seake-Kwawu A, Ghartey G, Asamoah YK, Siaw ADJ, et al. Hepatitis C virus seroprevalence, testing, and treatment capacity in public health facilities in Ghana, 2016–2021; A multi-centre cross-sectional study. Plos one. 2023;18(6):e0287580.

17. Duah E, Mathebula EM, Maluleke K, Azumah DE, Ephraim RKD, Mashamba-Thompson T. Evaluating performance of the Bioline™ HCV point-of-care test in Ghana. BMC Infectious Diseases. 2025;25(1):1327.

18. WHO. Renewed hope for all people living with viral hepatitis in Ghana 2023 [Available from: https://www.who.int/news-room/feature-stories/detail/renewed-hope-for-all-people-living-with-viral-hepatitis-in-ghana.

19. Hofmeister MG, Rosenthal EM, Barker LK, Rosenberg ES, Barranco MA, Hall EW, et al. Estimating Prevalence of Hepatitis C Virus Infection in the United States, 2013-2016. Hepatology. 2019;69(3):1020–31.

20. Rao VB, Johari N, du Cros P, Messina J, Ford N, Cooke GS. Hepatitis C seroprevalence and HIV co-infection in sub-Saharan Africa: a systematic review and meta-analysis. Lancet Infect Dis. 2015;15(7):819–24.

21. Okonkwo U, Okpara H, Otu A, Ameh S, Ogarekpe Y, Osim H, et al. Prevalence of hepatitis B, hepatitis C and human immunodeficiency viruses, and evaluation of risk factors for transmission: Report of a population screening in Nigeria. South African Medical Journal. 2017;107(4):346–51.

22. Fosu PK, Adjei CA, Atibila F, Aovare P, Ruiter RAC, Hoor GT. Prevalence of hepatitis C viral infection in Ghana: a systematic review and meta-analysis. BMC Infect Dis. 2025;25(1):1413.

23. Madison A, Adjei G, Agyeman S, Banson C, Nyane B, Arthur RA, et al. HBV and HCV prevalence and treatment eligibility assessment at a primary health facility in Ghana. medRxiv. 2024:2024.05.01.24306678.

24. Baber AS, Suganthan B, Ramasamy RP. Current advances in Hepatitis C diagnostics. Journal of Biological Engineering. 2024;18(1):48.

25. Dai S, Wang Z, Guo Q, Tang G, Guo Q, Zhang J, et al. Awareness of hepatitis C prevention and treatment and high-risk behaviors among the general population in Anhui Province: a cross-sectional study. Frontiers in Public Health. 2025;Volume 13 - 2025.

26. Adepoju VA, Udah DC, Ezenwa CA, Ganiyu J, Adnani QES. Awareness and Perception of Hepatitis C Self-Testing in Nigeria: A National Survey of Stakeholders and the Public. Venereology. 2024;3(4):199–210.

27. Adoba P, Boadu SK, Agbodzakey H, Somuah D, Ephraim RK, Odame EA. High prevalence of hepatitis B and poor knowledge on hepatitis B and C viral infections among barbers: a cross-sectional study of the Obuasi municipality, Ghana. BMC Public Health. 2015;15:1041.

28. Lee J, Di Ruggiero E. How does informal employment affect health and health equity? Emerging gaps in research from a scoping review and modified e-Delphi survey. Int J Equity Health. 2022;21(1):87.

29. WHO. Social determinants of health 2025 [Available from: https://www.who.int/news-room/fact-sheets/detail/social-determinants-of-health.

30. Zajacova A, Lawrence EM. The Relationship Between Education and Health: Reducing Disparities Through a Contextual Approach. Annu Rev Public Health. 2018;39:273–89.

31. Tachi K. Hepatitis C virus infection in Ghana: time for action is now. Ghana Medical Journal. 2018;52(1):1.

32. Tantuoyir MM, Camara M, Sohrabi M, SeyedAlinaghi S, Ahmadinejad Z. A rapid review of the epidemiology and combating strategies of hepatitis C virus infection in Ghana. Journal of Clinical Virology Plus. 2024;4(4):100195.

33. WHO. Hepatitis C 2025 [Available from: https://www.who.int/news-room/fact-sheets/detail/hepatitis-c.

34. Kuniholm MH, Jung M, Del Amo J, Talavera GA, Thyagarajan B, Hershow RC, et al. Awareness of Hepatitis C Virus Seropositivity and Chronic Infection in the Hispanic Community Health Study/Study of Latinos (HCHS/SOL). J Immigr Minor Health. 2016;18(6):1257–65.

35. Eberhardt MS, Pamuk ER. The importance of place of residence: examining health in rural and nonrural areas. Am J Public Health. 2004;94(10):1682–6.

36. Qaiser T, Bilal M, Shafiq M, Ahmed W, Asif M, Butt AM, et al. Investigating the role of social determinants in shaping health disparities: A public health lens on addressing inequities in urban and rural communities. European Chemical Bulletin. 2023;12(12):3843–9.

37. Saadati SM. The future of health equity: policy strategies to reduce disparities in public health. Journal of Foresight and Health Governance. 2025;2(2):15–32.

